# Mathematical Modeling and Analysis of COVID-19 pandemic in Nigeria

**DOI:** 10.1101/2020.05.22.20110387

**Authors:** Enahoro Iboi, Oluwaseun O. Sharomi, Calistus Ngonghala, Abba B. Gumel

## Abstract

A novel Coronavirus (COVID-19), caused by SARS-CoV-2, emerged from the Wuhan city of China at the end of 2019, causing devastating public health and socio-economic burden around the world. In the absence of a safe and effective vaccine or antiviral for use in humans, control and mitigation efforts against COVID-19 are focused on using non-pharmaceutical interventions (aimed at reducing community transmission of COVID-19), such as social (physical)-distancing, community lockdown, use of face masks in public, isolation and contact tracing of confirmed cases and quarantine of people suspected of being exposed to COVID-19. We developed a mathematical model for understanding the transmission dynamics and control of COVID-19 in Nigeria, one of the main epicenters of COVID-19 in Africa. Rigorous analysis of the Kermack-McKendrick-type compartmental epidemic model we developed, which takes the form of a deterministic system of nonlinear differential equations, reveal that the model has a continuum of disease-free equilibria which is locally-asymptotically stable whenever a certain epidemiological threshold, called the *control reproduction* (denoted by ℛ_*c*_), is less than unity. The epidemiological implication of this result is that the pandemic can be effectively controlled (or even eliminated) in Nigeria if the control strategies implemented can bring (and maintain) the epidemiological threshold (ℛ_*c*_) to a value less than unity. The model, which was parametrized using COVID-19 data published by Nigeria Centre for Disease Control (NCDC), was used to assess the community-wide impact of various control and mitigation strategies in the entire Nigerian nation, as well as in two states (Kano and Lagos) within the Nigerian federation and the Federal Capital Territory (FCT Abuja). It was shown that, for the worst-case scenario where social-distancing, lockdown and other community transmission reduction measures are not implemented, Nigeria would have recorded a devastatingly high COVID-19 mortality by April 2021 (in hundreds of thousands). It was, however, shown that COVID-19 can be effectively controlled using social-distancing measures provided its effectiveness level is at least moderate. Although the use of face masks in the public can significantly reduce COVID-19 in Nigeria, its use as a sole intervention strategy may fail to lead to the realistic elimination of the disease (since such elimination requires unrealistic high compliance in face mask usage in the public, in the range of 80% to 95%). COVID-19 elimination is feasible in both the entire Nigerian nation, and the States of Kano and Lagos, as well as the FCT, if the public face masks use strategy (using mask with moderate efficacy, and moderate compliance in its usage) is complemented with a social-distancing strategy. The lockdown measures implemented in Nigeria on March 30, 2020 need to be maintained for at least three to four months to lead to the effective containment of COVID-19 outbreaks in the country. Relaxing, or fully lifting, the lockdown measures sooner, in an effort to re-open the economy or the country, may trigger a deadly second wave of the pandemic.

## 1 Introduction

A novel Coronavirus (COVID-19), caused by SARS-CoV-2, emerged out of Wuhan city of China at the end of 2019. The pandemic, which has rapidly spread to over 210 countries, continues to inflict severe public health and socio-economic burden in many parts of the world, including in Nigeria. It has, as of May 18, 2020, accounted for over 4.7 million confirmed cases and about 315,000 deaths globally [1–3]. There is currently no safe and effective vaccine or antiviral for use against the pandemic in humans. Consequently, the control and mitigation efforts against the pandemic are focused on implementing non-pharmaceutical interventions (NPIs), such as social (physical)-distancing, community lockdown, contact tracing, quarantine of suspected cases, isolation of confirmed cases and the use of face masks in public [4–6]. Social-distancing (also referred to as physical-distancing) entails maintaining a physical distance of 2 metres (or 6 feet) from other humans in public gatherings. Community lockdown entails implementing the stay-at-home or stay-in-shelter strategy (so that people stay at, and work from, home), the closure of schools and non-essential businesses and services, avoiding large public or private gatherings etc.

Nigeria, the most populous country in Africa (with estimated population of 200 million [7]), is one of the epicenters of COVID-19 in Africa. It reportedly recorded its first COVID-19 case on February 27, 2020 (when an Italian citizen, who works in Nigeria, was diagnosed with the disease upon returning back to Nigeria from a trip to Milan, Italy [8]). This was the very first reported case in sub-Saharan Africa [8]. Data from the Nigeria Centre for Disease Control (NCDC) show that, as of May 18, 2020, Nigeria has 6,175 confirmed COVID-19 cases and 191 cumulative deaths (see Figure 1 for a time series data on confirmed COVID-19 cases in Nigeria)[9]. Furthermore, a total of 35,345 samples were tested for COVID-19 as of May 18, 2020 [9]. The federal government of Nigeria implemented a strict lockdown of three major centres in the country (namely, Lagos city, Ogun State and the Federal Capital Territory Abuja) on March 30, 2020, aimed at minimizing community transmission of COVID-19. During the imposed lockdown, only food stores and essential service providers were allowed to operate [10]. However, due to the increase in the cumulative COVID-19 confirmed cases, the government extended the community lockdown to include the rest of the country on April 27, 2020. The government announced plans to begin easing restrictions in the country from May 4, 2020, by allowing offices, businesses, markets, and stores to resume operation with limited hours and staff capacity, but with compulsory wearing of face masks in public and checking of body temperatures [10, 11]. However, restrictions, which include an overnight curfew and a ban on non-essential interstate travel, are still in place [10, 11]. A natural question to ask is when would it be safe to relax the lockdown measures to reopen the economy and the country? In other words, under what conditions can the lockdown measures be relaxed without risking the possibility of a second wave of COVID-19 that may be as (or even more) devastating as the first? This is one of the main objectives of the current study. We seek to use mathematical modeling approaches and rigorous mathematical analyses, coupled with statistical data analytics, to achieve this and related objectives.

**Figure 1:**
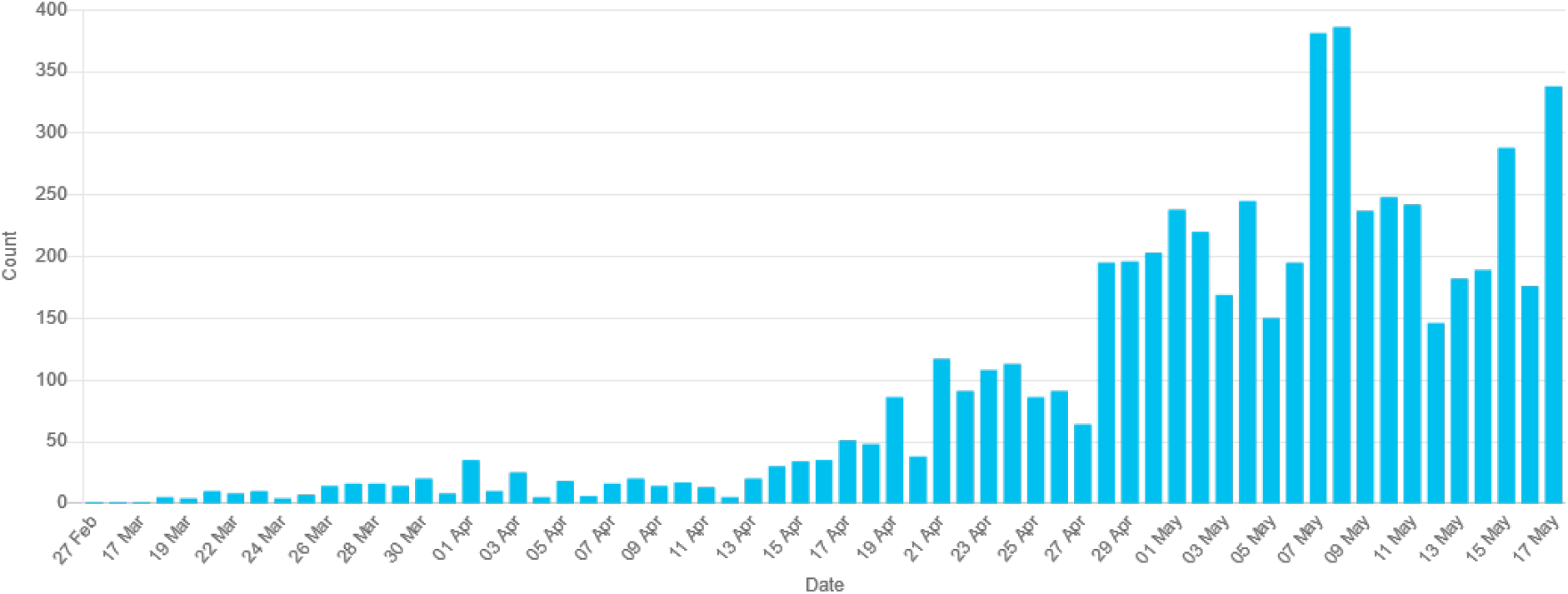
Confirmed COVID-19 Cases in Nigeria as at June 29, 2020. *Source: Nigeria Centre for Disease Control* [9]. Mathematical models have, historically, been used to provide realistic insight into the transmission dynamics and control of infectious diseases, dating back to the pioneering works of Sir Ronald Ross and Kermack-McKendrick in the 1900s [12]. Numerous models have also been designed and used to understand the mechanisms for the spread, control and mitigation of COVID-19 in a community. Ferguson *et al*. [13] developed one of the very first models for COVID-19. Their agent-based model, which was used to assess the impact of NPIs on COVID-19 mortality, predicted an alarmingly-high projection for the cumulative mortality in the US (2.2 million deaths) and the UK (510,000 deaths) if no public health interventions are used (i.e., their worst-case scenario predictions for COVID-induced mortality for the US was in millions, while that of the UK was in hundreds of thousands). A recent study by Eikenberry *et al*. [4] used a new multi-group Kermack-McKendrick-type epidemic model to assess the impact of mask usage in public in curtailing the spread of COVID-19 in the US. Their study shows that the use of face masks by members of the general public is potentially of high value in curtailing community transmission and the burden of the pandemic. This study further shows that the community-wide benefits of face masks are likely to be greatest when they are used in conjunction with other non-pharmaceutical interventions (such as social-distancing), and when face masks are adopted universally (i.e., throughout the nation) and compliance in their usage is high.

A detailed mathematical model for assessing the community-wide impact of NPIs on combating and mitigating the burden of COVID-19 was developed by Ngonghala *et al*. [5]. Their study showed that, while the early relaxation or lifting of social-distancing and community lockdown measures (and face mask usage in the public) is likely to lead to second wave, extended the duration of the social-distancing and lockdown measures (and face mask usage in public) can significantly reduce the COVID-induced mortality in the US in general, and the state of New York in particular. The potential for a COVID-19 outbreak aboard the Diamond Princess cruise (which experienced a major COVID-19 outbreak during the months of January and February of 2020) was modeled by Mizumoto and Chowell [14]. Their study showed a high estimate of the reproduction of the model (making major outbreak inevitable), and that the reproduction number substantially decreases with increasing effectiveness of the quarantine and isolation measures implemented on the ship.

Using a stochastic model, Hellewell *et al*. [15] showed that (for most instances) the spread of COVID-19 can be effectively contained in 3 months if contact-tracing and isolation are highly effective. Furthermore, using another stochastic model to study the COVID-19 trajectory in the Wuhan city of China from January to February, 2020, Kucharski *et al*. [16] showed that a reduction in COVID-19 transmission can be achieve when travel restrictions are implemented. Using a model for assessing the effect of mass influenza vaccination on the spread of COVID-19 and other influenza-like pathogens co-circulating during an influenza season, Li *et al*. [17] showed that increasing influenza vaccine uptake (or enhancing the public health interventions) would facilitate the management of outbreaks of respiratory pathogens circulating during the peak flu season.

Recently, Iboi *et al*. [6] developed a mathematical model to determine whether or not a hypothetical imperfect vaccine can lead to the elimination of COVID-19 in the United States. Their study showed that such elimination is feasible, using the hypothetical vaccine with assumed efficacy of 80%, if the vaccine coverage is high enough to achieve herd immunity. In particular, the vaccine coverage needed to achieve herd immunity in US is 90%, while the computed herd threshold for the states of New York and the state of Florida are 84% and 85%, respectively.

The current study is based on using a mathematical model to assess the impact of NPIs on the transmission dynamics of the COVID-19 pandemic in Nigeria. The model will be parametrized using available COVID-19 mortality data for Nigeria, the state of Lagos, Kano and the Federal Capital Territory Abuja to estimate important parameters related to the reduction in community contacts. The main objectives of the study include determining whether or not the current NPI-based control and mitigation measures in Nigeria would be adequate to lead to the effective control of the pandemic in Nigeria. Further, the impact of early relaxation or lifting of the current social-distancing and community lockdown measures will be assessed. In particular, the model will be used to determine when it would be safe to relax these measures without risking the possibility of a second wave of the pandemic.

## 2 Materials and Methods

### 2.1 Model Formulation

The model is developed by splitting the total human population at time *t*, denoted by *N* (*t*), into the mutually-exclusive compartments of susceptible (*S*(*t*)), exposed (*E*(*t*)), symptomatically-infectious (*I*_*s*_(*t*), asymptomatically-infectious (*I*_*a*_(*t*)), hospitalized (*I*_*h*_(*t*)) and recovered (*R*(*t*)) individuals. Thus,

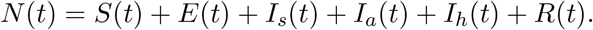

It should be emphasized that the epidemiological compartment *I*_*a*_ also contain individuals who may show mild symptoms of the disease. Furthermore, the compartment *I*_*h*_ for hospitalization also includes those with clinical symptoms of COVID-19 who are self-isolating at home. The model is given by the following deterministic system of nonlinear differential equations (where a dot represents differentiation with respect to time *t*):

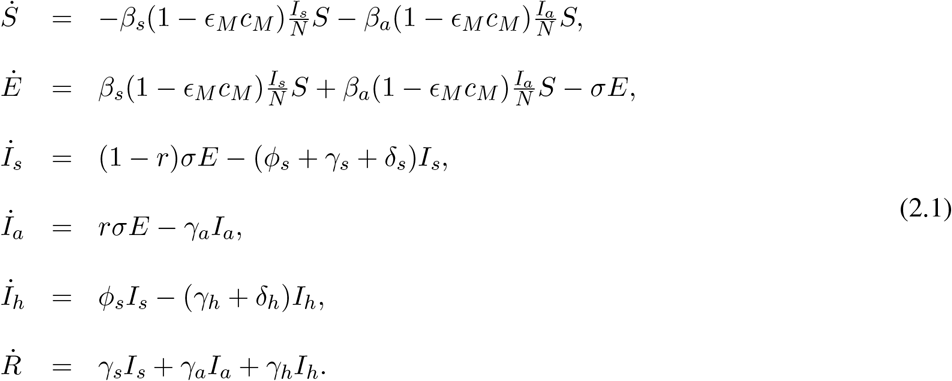

A flow diagram of the model is depicted in Figure 2. The state variables and parameters of the model are described in Tables 1 and 2, respectively.

**Figure 2:**
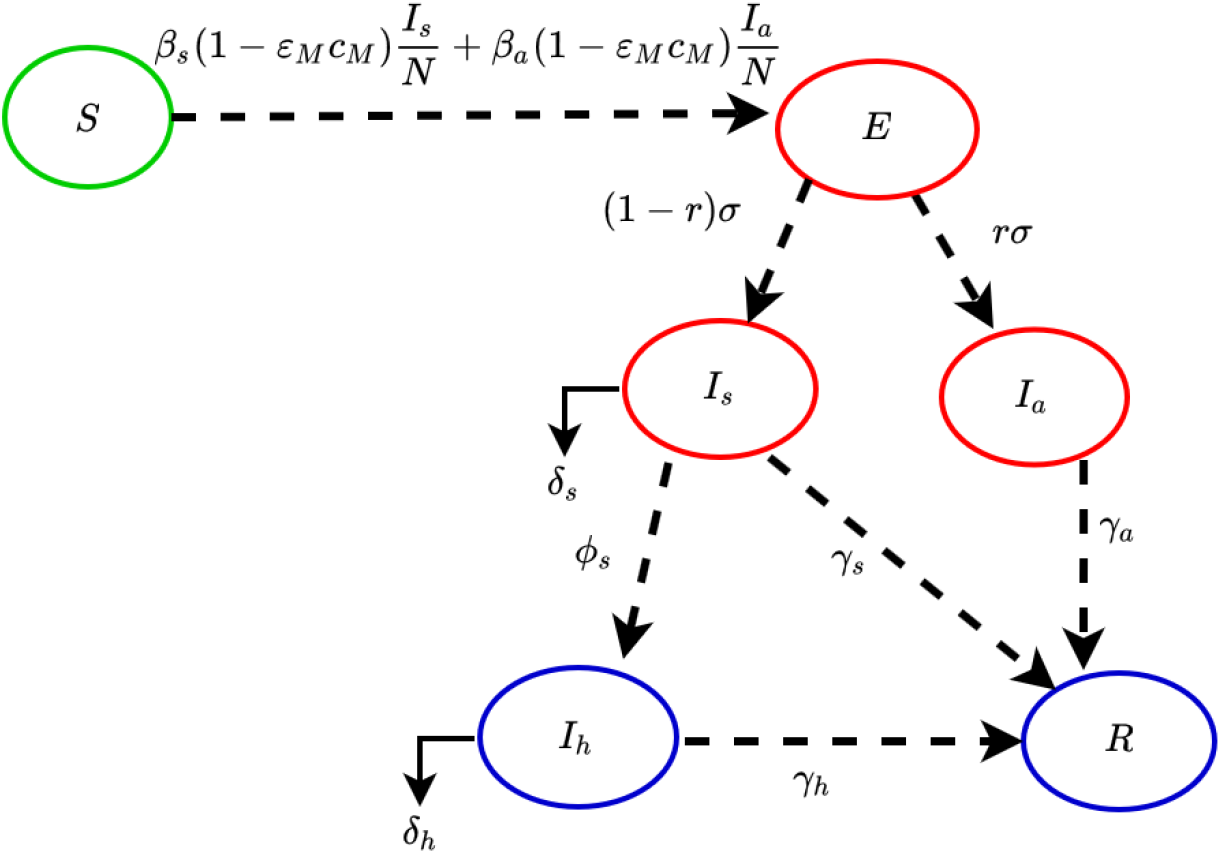
Flow diagram of the COVID-19 model (2.1).

**Table 1:** Description of state variables of the COVID-19 model (2.1).

| State variable | Description |
| --- | --- |
| $S$ | Population of susceptible individuals |
| $E$ | Population of exposed individuals |
| $I_s$ | Population of symptomatically-infectious individuals |
| $I_a$ | Population of asymptomatically-infectious individuals |
| $I_h$ | Population of hospitalized individuals |
| $R$ | Population of recovered individuals |

**Table 2:** Description of the parameters of the COVID-19 model (2.1).

| Parameter | Description |
| --- | --- |
| $\beta_s(\beta_a)$ | Effective community contact rate (a measure of social-distancing effectiveness) |
| $c_M$ | Proportion of members of public who wear masks in public (i.e., masks compliance) |
| $\epsilon_M$ | Efficacy of face-masks to prevent acquisition of infection by susceptible individuals |
| $\sigma$ | Rate of progression from the exposed compartment to the infectious compartment ($1/\sigma$ is the incubation period) |
| $1 - r$ | Fraction of exposed individuals who show clinical symptoms at the end of the incubation period |
| $\gamma_s(\gamma_a)(\gamma_h)$ | Recovery rate for individuals in the $I_s(I_a)(I_h)$ compartment |
| $\delta_s(\delta_h)$ | Disease-induced mortality rate for infectious (hospitalized) individuals |
| $\phi_s$ | Hospitalization rate for infectious individuals |

In the model (2.1), the parameter *β*_*s*_(*β*_*a*_) represents the effective contact rate (i.e., contacts capable of leading to COVID-19 transmission) for symptomatically-infectious (asymptomatically-infectious) individuals, 0 *< c*_*M*_ ≤ 1 is the proportion of members of the public who wear face-masks (correctly and consistently) in public and 0 *< ϵ*_*M*_ *≤* 1 is the efficacy of the face-masks. This formulation (i.e., *β*_*s*_ ≠ *β*_*a*_) allows us to account for the possible heterogeneity in the contact rates of infectious with (*I*_*s*_) or without (*I*_*a*_) clinical symptoms of COVID-19. The parameter *σ* represents the progression rate of exposed individuals (i.e., 1*/σ* is the intrinsic incubation period of COVID-19). A proportion, 0 *<r* ≤ 1, of exposed individuals show no clinical symptoms of COVID-19 (and move to the class *I*_*a*_) at the end of the incubation period. The remaining proportion, 1 − *r*, show clinical symptoms and move to the *I*_*s*_ class. The parameter *γ*_*s*_(*γ*_*a*_)(*γ*_*h*_) represents the recovery rate for individuals in the *I*_*s*_(*I*_*a*_)(*I*_*h*_) class. Similarly, *ϕ*_*s*_ is the hospitalization (or self-isolation) rate of individuals with clinical symptoms of COVID-19. Finally, the parameter *δ*_*s*_(*δ*_*h*_) represents the COVID-induced mortality rate for individuals in the *I*_*s*_(*I*_*h*_) class.

It is convenient, for housekeeping purposes, to define the state variable *D*(*t*) for the number of COVID-deceased individuals, to keep track of COVID-19 related deaths. It then follows from some of the equations of the model (2.1) that the rate of change of the population of COVID-deceased individuals is given by:

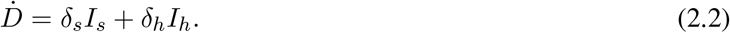

### 2.2 Data Fitting and Parameter Estimation

Estimates for some of the parameters of the model (2.1) are available from the literature (as indicated in Table 4). Other parameters, such as the community contact rate parameters (*β*_*a*_ and *β*_*s*_), the hospitalization rate (*ϕ*_*s*_) and the COVID-induced mortality parameters (*δ*_*h*_ and *δ*_*s*_), are unknown. Consequently, we use the data published by the NCDC [9] to estimate these unknown parameters for the entire Nigerian nation, the States of Kano and Lagos and the FCT. Furthermore, we estimate the respective basic reproduction number (ℛ_0_) for each of the four communities. In particular, we fit the cumulative number of deaths generated from the model (2.1) to the observed cumulative COVID-19 mortality data for the four communities reported by the NCDC [9]. The data-fitting process involves implementing the Markov Chain Monte Carlo (MCMC) sampling method in the statistical software package R. This process produces 50,000 different parameter sets that are now used with a nonlinear least squares method to estimate the values of the aforementioned unknown parameters as well as the associated 95% confidence intervals (CIs). The nonlinear least squares method allows the determination of the set of parameters that minimizes the sum of the squares of the differences between the cumulative deaths predicted by the model (2.1) and the observed cumulative deaths (for the entire Nigerian nation, the two states of Kano and Lagos, and the Federal Capital Territory, Abuja). The estimated values of the unknown parameters and the basic reproduction numbers, and their 95% confidence intervals, are tabulated in Table 3. Figure 3 depicts the fitting of the observed and predicted cumulative deaths for the four communities.

**Table 3:** Fitted parameters of the model (2.1) using COVID-19 cumulative case data for (a) Nigeria (b) Lagos State (c) Kano State (d) FCT Abuja. Cumulative case data obtained from NCDC [9].

| Fitted parameters | Nigeria | Lagos State | Kano State | FCT Abuja |
| --- | --- | --- | --- | --- |
| $\beta_s$ | 1.082 [0.855, 1.1] | 0.606 [0.589, 0.606] | 0.419 [0.379, 0.433] | 0.413 [0.413, 0.459] |
| $\beta_a$ | 0.302 [0.302, 0.344] | 0.334 [0.331, 0.334] | 0.480 [0.432, 0.768] | 0.382 [0.317, 0.382] |
| $\delta_s$ | 0.288 [0.243, 0.291] | 0.133 [0.118, 0.133] | 0.205 [0.185, 0.308] | 0.286 [0.286, 0.291] |
| $\delta_h$ | 0.151 [0.151, 0.238] | 0.065 [0.065, 0.139] | 0.272 [0.272, 0.435] | 0.254 [0.055, 0.254] |
| $\phi_s$ | 0.154 [0.069, 0.183] | 0.151 [0.117, 0.151] | 0.438 [0.394, 0.438] | 0.182 [0.119, 0.182] |
| $\mathcal{R}_0$ | 1.98 [1.98, 2.097] | 1.878 [1.878, 1.94] | 1.948 [1.753, 1.948] | 1.675 [1.524, 1.675] |

**Table 4:** Default (baseline) values of the parameters of the model (2.1).

| Parameter | Default (baseline) value | Reference |
| --- | --- | --- |
| $\epsilon_M$ | 0.5 (dimensionless) | Estimated from [4, 18] |
| $c_M$ | 0.1 (dimensionless) | Estimated from [4, 5] |
| $\sigma$ | 1/6 day<sup>-1</sup> | [19, 20] |
| $r$ | 0.35 (dimensionless) | [13, 20–22] |
| $\gamma_a$ | 1/7 day<sup>-1</sup> | [23, 24] |
| $\gamma_s$ | 1/7 day<sup>-1</sup> | [23, 24] |
| $\gamma_h$ | 1/14 day<sup>-1</sup> | [23, 24] |

**Figure 3:**
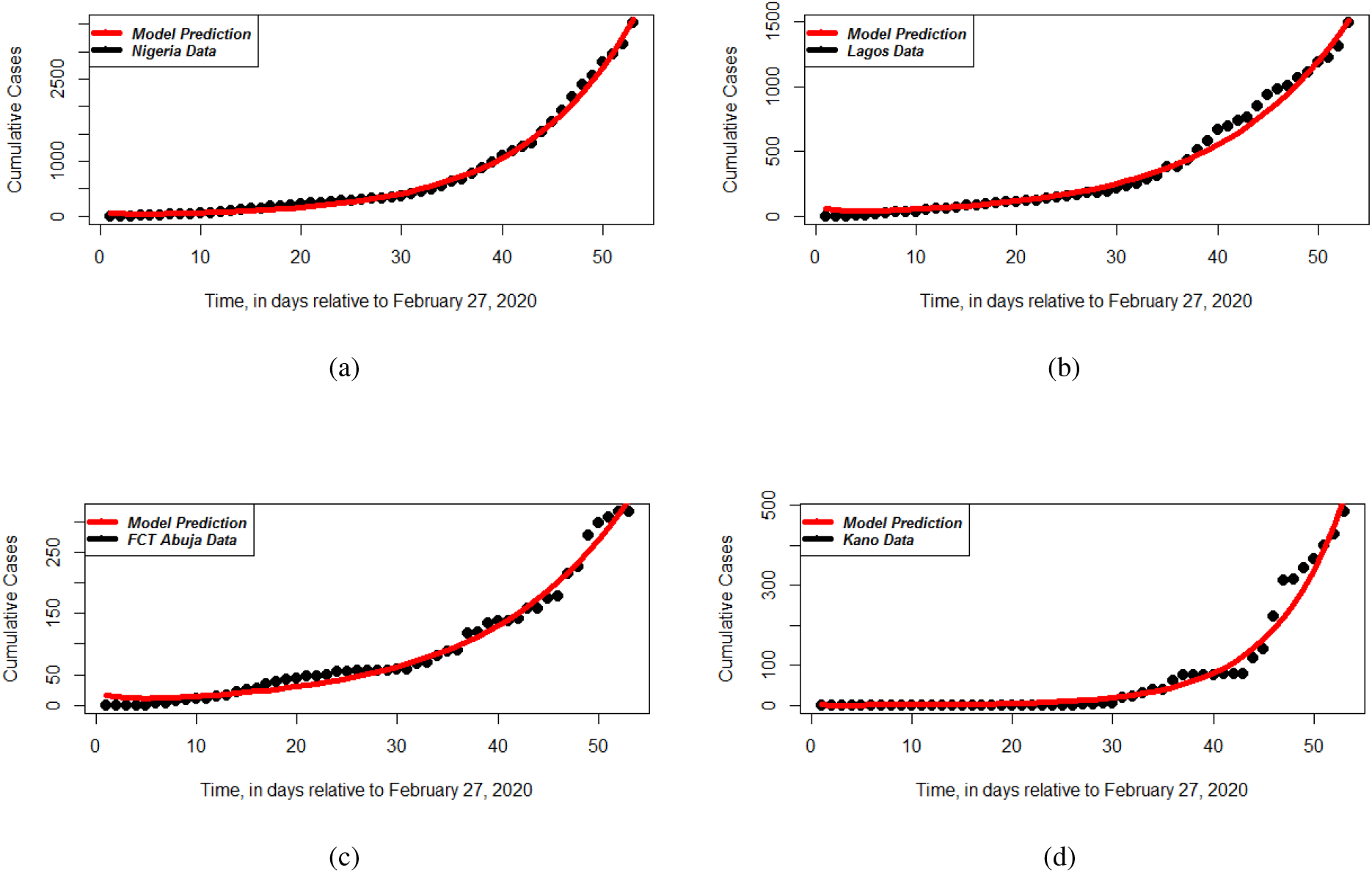
Data fitting of the model (2.1) using COVID-19 cumulative case data for (a) Nigeria (b) Lagos State (c) Kano State (d) FCT Abuja. Cumulative case data obtained from NCDC [9].

## 3 Results

### 3.1 Analytical Results: Asymptotic Stability Analysis of Continuum of Disease-free Equilibria

The model (2.1) has a continuum of disease-free equilibria (DFE), given by

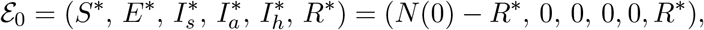

where *N* (0) is the initial total sizes of the population and 0 *≤ R*^*∗*^ *≤ N* (0). The next generation operator method [25, 26] can be used to analyse the asymptotic stability property of the DFE. In particular, using the notation in [25], it follows that the associated next generation matrices, *F* and *V*, for the new infection terms and the transition terms, are given, respectively, by

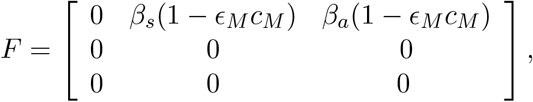

and,

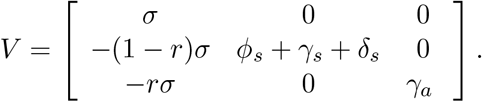

It is convenient to define the quantity *ℛ*_*c*_ by:

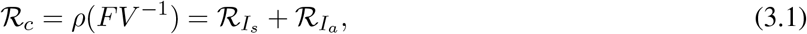

where *ρ* is the spectral radius,

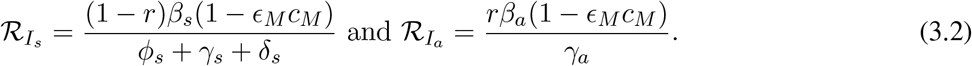

The quantity ℛ_*c*_ is the *control reproduction number* of the model (2.1). It measures the average number of new COVID-19 cases generated by a typical infectious individual introduced into a population where a certain fraction is protected. It is the sum of the constituent reproduction numbers associated with the number of new cases generated by symptomatically-infectious humans 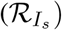 and asymptomatically-infectious 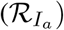 individuals. The reproduction number *ℛ*_*c*_ will now be interpreted epidemiologically below.

#### 3.1.1 Interpretation of the reproduction number of the model

As stated above, the reproduction number *ℛ*_*c*_ is the sum of the two constituent reproduction numbers, 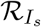 and 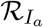. The constituent reproduction number 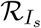 is given by the product of the infection rate of susceptible individuals by symptomatically-infectious humans, near the DFE *β*_*s*_(1 *−ϵ* _*M*_ *c*_*M*_), the proportion of exposed individuals that survived the incubation period and moved to the symptomatically-infectious class 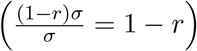 and the average duration in the asymptomatically-infectious class 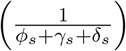. Similarly, the constituent reproduction number 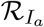 is given by the product of the infection rate of susceptible individuals, near the DFE *β*_*a*_(1 *−ϵ* _*M*_ *c*_*M*_), the proportion that survived the exposed class and moved to the asymptomatically-infectious class 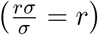 and the average duration in the *I*_*a*_ class 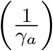. The sum of 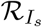 and 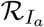 gives *ℛ*_*v*_.

The result below follows from Theorem 2 of [25].

**Theorem 3**.**1**. *The disease-free equilibrium (DFE) of the model* (2.1) *is locally-asymptotically stable if ℛ*_*c*_ *<* 1. *If ℛ*_*c*_ *>* 1, *the epidemic grows rapidly, reaches a peak, and eventually declines to zero*.

Theorem 3.1 can be interpreted, epidemiologically, to mean that a small influx of COVID-19 cases will not generate a COVID-19 outbreak if the control reproduction number (ℛ_*c*_) is less than unity. It should be mentioned that, for epidemic models such as (2.1), the epidemiological requirement ℛ_*c*_ *<* 1 is only sufficient, but not necessary, for curtailing or eliminating the epidemic. This is owing to the fact that, for epidemic models (i.e., models with no vital/demographic dynamics, such as (2.1)), the epidemic always dies out with time (regardless of the value of the reproduction number of the model). In other words, even if the value of the reproduction number (ℛ_*c*_) exceeds unity, the epidemic will eventually die out. This is because (if ℛ_*c*_ *>* 1), a combination of factors, such as the implementation of control measures in the community and the increasing level of natural infection-acquired immunity in the community, will greatly curtail the pandemic to the extent that sustained community transmission is not feasible (and the epidemic eventually dies out). It should be recalled that one of the main assumptions in the formulation of the model (2.1) is that recovered individuals acquire permanent immunity against future COVID-19 infection. The consequence of this natural immunity build up in the community is that many susceptible members of the community are now being protected from acquiring COVID-19 infection. It should, however, be mentioned that, for endemic models (where the aforementioned vital/demographic dynamics are allowed), the epidemic will persist in the community whenever ℛ_*c*_ *>* 1. The reason for this is that, by allowing for the recruitment of susceptible individuals (by birth or immigration), the population of wholly-susceptible (i.e., COVID-naive) individuals is continually being replenished, thereby allowing the disease to find potential targets to infect. This allows the epidemic to sustain itself in the community.

It is convenient to define the following feasible (positively-invariant and attracting) region for the model (2.1)

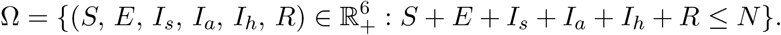

We claim the following result:

**Theorem 3**.**2**. *The continuum disease-free equilibria (ε*_0_*) of the model* (2.1) *is globally-asymptotically stable in* Ω *if ℛ*_*c*_ *≤* 1.

The proof to Theorem 3.2, based on using Lyapunov function theory, is given in Appendix A. Here, too, the requirement ℛ_*c*_ *≤* 1 is sufficient, and not necessary, for the elimination of the pandemic. The epidemiological implication of this result is that COVID-19 elimination is independent of the initial sizes of the sub-populations of the model (i.e., the initial number of infected individuals introduced into the community does not have to be within the basin of attraction of the continuum of the disease-free equilibria, ε _0_ of the model).

Using the baseline and fitted parameter values in Tables 3 and 4, the value of the control reproduction number (ℛ_*c*_) for Nigeria, the states of Lagos and Kano, as well as the Federal Capital Territory Abuja, are, respectively, ℛ_*c*_ = 2.13, 1.91, 2.55 and ℛ_*c*_ = 1.81. In the absence of any public health intervention implemented in Nigeria, the control reproduction number becomes the *basic reproduction number* [12], given by

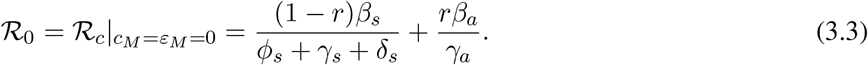

The quantity ℛ_0_ represents the average number of secondary infections generated by a typical infected individual introduced into a completely susceptible population during the duration of the infectiousness of the individual. The fitted values of *ℛ*_0_ for Nigeria, the states of Kano and Lagos, as well as the FCT Abuja, are tabulated in Table 3. It follows, from the values of ℛ_*c*_ above (for Nigeria and the States of Kano and Lagos and the FCT) and the fitted values of ℛ_0_ in Table 3, that the control strategies adopted in Nigeria (and in the two states and territory) have only marginally reduced the value of the basic reproduction number ℛ_0_. Further, in each of the four communities, the value of the control reproduction number exceeds unity. Hence, the pandemic will persist in all four communities (since ℛ_*c*_ need to be brought to a value less than unity to have any chance to effectively control or eliminate the pandemic, in line with Theorem 3.1). In other words, the current levels of NPIs implemented in the country are inadequate to lead to the effective control (or elimination) of the pandemic in Nigeria (and in any of the two states and territory). Thus, this study clearly shows that the effectiveness and coverage levels of the currently-adopted public health interventions must be significantly enhanced to enable such effective control or elimination of the pandemic.

It should be noted from the estimated (fitted) values of the basic reproduction number of the model (2.1) in Table 3 that, during the early stage of the pandemic, a typical COVID-19 infected individual in Nigeria was transmitting, on average, to two other people in Nigeria. In other words, the pandemic was spreading at this exponential rate between February 27, 2020 until the time control and mitigation measures were implemented (i.e., until March 30, 2020, when the government of Nigeria announced lockdown measures in some localities). This certainly suggests that a large number of Nigerians were already infected with COVID-19 before the lockdown measures were implemented. This also implies that a large number of Nigerians may have already died of COVID-19 during this period of explosive growth of the pandemic (particularly the elderly and those with underlying health conditions). The COVID-19 case and mortality numbers were obviously not recorded in Nigeria during the February 27, 2020 to March 30, 2020 time period when the public health system did not really kick in. This basic fact clearly suggests that, even in the best case scenario when in fact the index case was indeed recorded on the reported February 27, 2020 date, Nigeria was at least a month behind the COVID-19 pandemic. This certainly is a mountain distance to climb to try to catch-up with the pandemic.

It is certainly reasonable to question the veracity of the claim that the pandemic was indeed brought to Nigeria on February 27, 2020. There are many reasons to support this scepticism. Nigeria has a major trading partnership with China. The volume of traffic between the two countries is enormous (involving people travelling back-and-forth for business, students going to, and returning from, China, people embarking on medical tourism, diplomatic relations etc.). Owing to the fact that many have claimed the pandemic was already spreading in China a month or two before the December 31, 2020 declaration by the Wuhan city public health authorities [5], it is certainly reasonable to consider the possibility of the introduction of COVID-19 to Nigeria by as early as the end of 2019. Further, the pandemic in China moved rapidly to Europe, and the fact that Nigeria has equally large volume of traffic to, and from, Europe also provides supporting evidence to the hypothesis that the pandemic may well have been circulating in Nigeria by the end of 2019. Similar situation occurred in New York city, where recent modeling data from Northeastern University in Boston showed that New York city might have had about 11,000 COVID-19 cases before the first confirmed case in the State was announced on March 1, 2020 [27]. The study, backed by CDC testing data at JFK International Airport, claimed that COVID-19 was already circulating in the State of New York since late January 2020. Considering the fact that some of Nigeria’s largest cities, notably Kano and Lagos, are very much like New York city in terms of both population size (with each city having at least 10 million inhabitants) and population density. Hence, it stands to reason that what transpired in New York city, *vis-a-vis* the true burden of the COVID-19 pandemic, should, in theory, be expected to transpire in these densely-populated Nigerian cities (considering also the healthcare capacity challenges in Nigeria). Our claim is simply that the COVID-19 data reported by the NCDC may be grossly under-estimating the true burden of the pandemic in Nigeria. Recent evidence of “mysterious deaths” in Kano seems to provide some justification to our claim. Further, it is quite reasonable to assume that a sizable proportion of Nigerians may have acquired the COVID-19 infection during the exponential growth phase of the outbreak (between February, 2020 to March, 2020) and may have developed natural immunity to it. This may, to a small extent perhaps, lay some credence to the remarkably low case and mortality COVID-19 numbers reported by the NCDC. However, only a robust serology testing, which is reportedly currently not been implemented in the nation, will provide a true measure of the natural herd immunity against COVID-19 in Nigeria.

It should also be observed from Table 3 that the fitted value of the basic reproduction number for COVID-19 in the Federal Capital Territory (FCT) is lower than those for Kano and Lagos States (as well as for the entire Nigerian nation). Thus, the FCT is expected to suffer lower COVID-19 burden than the two states (and the rest of the nation). The lower value of the basic reproduction number of the FCT can be intuitively explained based on the fact that the FCT is not as densely-populated as Kano or Lagos cities (or States).

### 3.2 Computation of Final Size of the Pandemic

In this section, the final size relation for the COVID-19 pandemic will be rigorously derived. Using the notation in [28], let 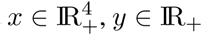, and *z* IR_+_ represent the sets of infected, susceptible and recovered components of the model. Thus,it follows from the model (2.1), that *x*(*t*)= (*E*(*t*), *I*_*s*_(*t*), *I*_*a*_(*t*), *I*_*h*_(*t*))^*T*^, *y*(*t*)= *S*(*t*) and *z*(*t*)= *R*(*t*). Further, following Arino *et al*. [28], let *D* be the *m× m* diagonal matrix whose diagonal entries, denoted by *σ*_*i*_(*i* = 1, 2, *…, m*), are the relative susceptibilities of the corresponding susceptible class. It is convenient to define Π to be an *n× n* matrix with the property that the (*i, j*) entry represents the fraction of the *j*^th^ susceptible compartment that goes into the *i*^th^ infected compartment upon becoming infected. Let *b* be an *n* dimensional row vector of relative horizontal transmissions. Using the notation in [28], let the infection rate, *λ*, of the model (2.1) be represented by *β*. That is, *λ* = *β*(*x, y, z*). Define the *m* dimensional vector Γ = [Γ_1_, Γ_2_, *…*, Γ_*m*_] = *βbV* ^*−*1^Π*D* [28]. It follows, in the context of the model (2.1), that

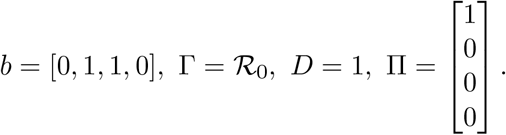

Using the above change and variables and definitions, the model (2.1) reduces to:

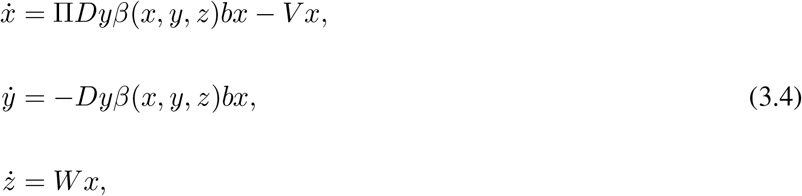

where *W* is a *k× n* matrix with the property that the (*i, j*) entry represents the rate at which individuals of the *j*^th^ infected compartment transition into the recovered (*i*^th^ *z*) compartment upon recovery and the matrix *V* is as defined in Section 3.1. It is worth stating that the reproduction number (*ℛ*_*c*_, of the model (2.1) (or, equivalently, (3.4)), can be recovered using the definition *ℛ*_*c*_ = *β*(0, *y*_0_, *z*_0_)*bV* ^*−*1^Π*Dy*_0_ given in Theorem 2.1 of [28] (it should be noted that this theorem also allows for recovering the local asymptotic stability result for the family of disease-free equilibria of the model (2.1), given in Section 3.1). Furthermore, the results below, for the final size relations of the model (2.1) (or, equivalently, (3.4)), can be established using Theorem 5.1 of [28].

**Theorem 3**.**3**. *Consider the epidemic model* (2.1) *(or, equivalently*, (3.4)*). The final size relations are given by*

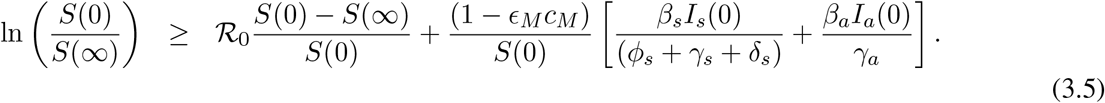

It is worth mentioning that, by setting *I*_*a*_(0) = 0, with *S*(0) *>* 0 and *I*_*s*_(0) *>* 0, the final size relations, given by the inequalities in (3.5), reduce to:

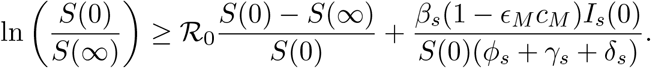

## 4 Numerical Simulations: Assessment of Control Strategies

The model (2.1) is now simulated, using the baseline parameter values tabulated in Tables 3 and 4, to assess the community-wide impact of the various intervention strategies incorporated into the model (namely social-distancing/community lockdown and the use of face masks in public) for the entire Nigerian nation in general, and the states of Lagos, Kano and the Federal Capital Territory (Abuja), in particular. For simulation convenience (and for computational tractability), we re-scale the total Nigerian population to 1.5 million (i.e., we set *S*_*u*_(0) = 1.5 million for Nigeria). Further, we re-scale the populations of each of the three states and the FCT to 1 million each (i.e., we set *S*_*u*_(0) = 1 million for each of the three states and the FCT in the numerical simulations). We simulate the pandemic starting with one index case in the symptomatically-infectious class (i.e., we set *I*_*s*_(0) = 1), while initially fixing all other state variables for the infected and recovered classes of the model to zero (i.e., we also set *E*(0) = *I*_*a*_(0) = *I*_*h*_(0) = *R*(0) = 0).

### 4.1 Effect of Social-distancing

The effect of social-distancing on the dynamics of COVID-19 is monitored by simulating the model, using the baseline parameter values tabulated in Tables 3 and 4 (with baseline mask compliance and efficacy), for various effectiveness levels of social-distancing. For simulation purposes, the following three effectiveness levels of social-distancing are considered (it should be mentioned that, the case with no social-distancing is equivalent to using the baseline values of the community contact rate parameters, *β*_*s*_ and *β*_*a*_, given in Table 3):

i. **Mild effectiveness level of social-distancing:** This entails reducing the community contact rates (*β*_*s*_ and *β*_*a*_) by 10% from their baseline values in Table 3. That is, *β*_*s*_ = 0.6597 (Nigeria), 0.7648 (Lagos State), 0.1832 (Kano State) and 0.2520 (FCT Abuja) *per* day. Similarly, *β*_*a*_ = 0.0913 (Nigeria), 0.1599 (Lagos State), 1.3390 (Kano State) and 0.5913 (FCT Abuja) *per* day.
ii. **Moderate effectiveness level of social-distancing:** This entails reducing the community contact rates (*β*_*s*_ and *β*_*a*_) by 30% from their baseline values in Table 3. That is, *β*_*s*_ = 0.5131 (Nigeria), 0.5949 (Lagos State), 0.1425 (Kano State), 0.1960 (FCT Abuja) *per* day. Similarly, *β*_*a*_ = 0.0710 (Nigeria), 0.1244 (Lagos State), 1.0415 (Kano State) and 0.4599 (FCT Abuja) *per* day.
iii. **Strict effectiveness level of social-distancing:** This entails reducing the community contact rates (*β*_*s*_ and *β*_*a*_) by 50% from their baseline values in Table 3. That is, *β*_*s*_ = 0.3665 (Nigeria), 0.4249 (Lagos State), 0.1018 (Kano State), 0.1400 (FCT Abuja) *per* day, and *β*_*a*_ = 0.0507 (Nigeria), 0.0889 (Lagos State), 0.7439 (Kano State), 0.3285 (FCT Abuja) *per* day.

Figure 4 depicts the daily cases and cumulative COVID-19 mortality for the four communities, for various levels of social-distancing effectiveness. In particular, Figure 4 (a) (blue curve) shows that, when no social-distancing measures are implemented in the country (i.e., when we simulate the worst-case scenario), Nigeria will attain a peak in the daily number of cases (of about 4,800) around July 28, 2020. This corresponds to a peak cumulative mortality of about 225,000, which is attained by February 2021 (Figure 4 (b), blue curve). It should be mentioned that this estimate for the cumulative COVID-19 mortality for Nigeria is a lot higher than what one would expect using the current mortality figures reported by the NCDC. There are many reasons to explain this major discrepancy. First of all, our cumulative mortality projections are for the worst-case scenario where the pandemic is spreading in the country and no public health interventions (such as social-distancing, community lockdown, quarantine and isolation measures etc.) are implemented in the nation. Hence, it is certainly intuitive that, in the absence of any public health intervention measures, the COVID-19 pandemic will be spreading exponentially fast, causing such unprecedented level of cases and mortality [5, 13]. The so-called “mysterious deaths” of hundreds of people in Kano during the month April, 2020 provide some justification to this claim. Secondly, the number of COVID-19 tests carried out in Nigeria is infinitesimally small. Data from the NCDC shows that only 35, 345 COVID-19 tests have been carried out in Nigeria as of May 18, 2020 [9]. Thirdly, no real autopsy or testing of deceased individuals is (universally) carried out in Nigeria. This is particularly more pronounced in the northern part of the country (and in some other faith communities in the southern part of the country), where deceased persons are buried within hours of death (and no autopsy or testing is carried out to determine the cause of death). Taking these factors in totality, it seems reasonably plausible to assume that the COVID-19 data provided by the NCDC represents a gross under-representation of the actual data. In other words, in the absence of sizable percentage of COVID-19 diagnostic testing conducted in Nigeria, the NCDC data most likely represents a gross under-estimation of the real burden of the COVID-19 pandemic in Nigeria. Hence, the predictions of reasonably-realistic mathematical models, such as the one we formulated in this study, may represent far more reliable representation of the true burden of the pandemic than the NCDC data portrays. Another point to emphasize is the possibility that the pandemic may well be spreading in Nigeria a lot earlier than the suggested February 27, 2020 date of initial introduction (i.e., index case), as stated in Section 3.1.1.

**Figure 4:**
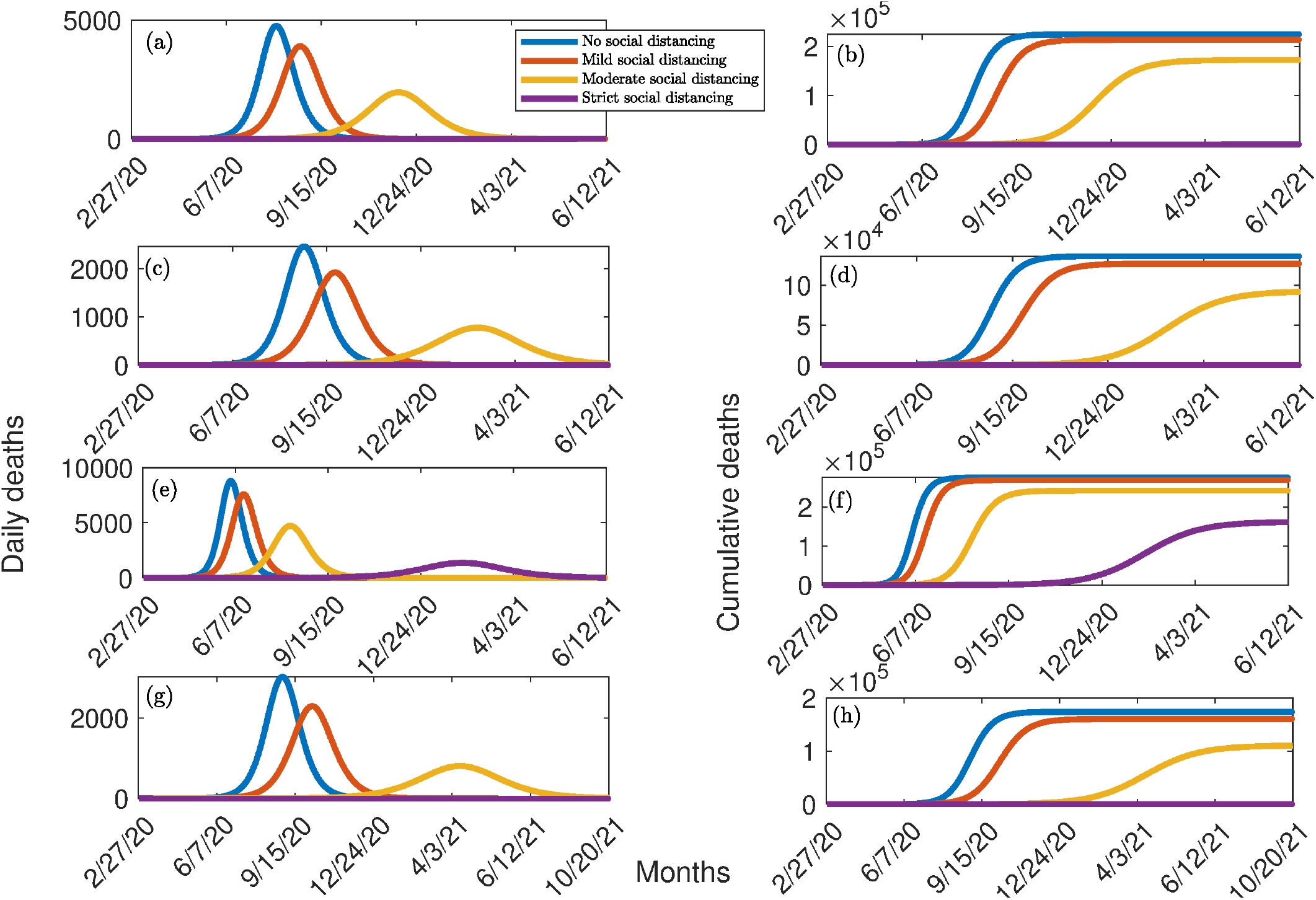
Simulations of the model (2.1) for various effectiveness of social-distancing control measures in the absence of mask usage. Mild social-distancing corresponds to 10% decrease in the values of *β*_*a*_ and *β*_*s*_ from their baseline values. Moderate social-distancing corresponds to 30% decrease in the values of *β*_*a*_ and *β*_*s*_ from their baseline values. Strict (high) social-distancing corresponds to 50% decrease in the values of *β*_*a*_ and *β*_*s*_ from their baseline values.(a) Daily cases for Nigeria, (b) cumulative deaths for Nigeria, (c) Daily cases for Lagos State, (d) cumulative deaths for Lagos State (e) Daily cases for Kano State, (f) cumulative deaths for Kano State, (g) Daily cases for FCT Abuja, (h) cumulative deaths for FCT Abuja. Parameter values used are as given in Tables 3 and 4.

We now simulate the model (2.1) to assess the impact of social-distancing on COVID-19 dynamics in Nigeria. Our simulations show that when mild social-distancing measures are implemented in the nation, the pandemic peak shifts to August 23, 2020, and the peak daily case decreased to about 3,900 (representing a 19% reduction from the peak daily cases corresponding to the worst-case scenario described above) (Figure 4 (a), red curve). The cumulative mortality, for this setting, reduced by 5%, to 213,500 (Figure 4 (b), red curve). Thus, while the implementation of mild social-distancing measures in Nigeria significantly decreases peak daily cases (by about 19%), it only marginally reduces COVID-19 mortality.

More dramatic reductions are observed when moderate social-distancing measures are implemented (Figures 4 (a) and (b), gold curve). In particular, the peak daily cases shifts to January 2021, with a peak daily case of 2000 (representing a whopping 58% reduction in the peak daily case, in comparison to the worst-case scenario baseline). Furthermore, the peak cumulative mortality reduced to 171,800 (attained around April 2021). This represents a 24% reduction in cumulative mortality nationwide. Thus, a dramatic reduction in COVID-19 burden will be achieved in Nigeria if moderate levels of social-distancing measures are implemented nationwide (with projected number of COVID-induced mortality not exceeding 171,800). If strict social-distancing measures were implemented (and maintained) during the early stage of the pandemic, our simulations show that the pandemic will fail to take off in the country (Figures 4 (a) and (b), purple curves…. which are superimposed with the x-axis). Thus, this study shows that COVID-19 can be effectively curtailed in Nigeria using moderate or strict social-distancing measures. In particular, COVID-19 would have had no chance to thrive in Nigeria if strict social-distancing measures were implemented early during the pandemic (and maintained for a certain period of time).

Simulations for Lagos State, for the worst-case scenario (with no social-distancing), show a peak daily case of about 2,500, attained by around August 2020 (Figure 4 (a), blue curve). This corresponds to a cumulative mortality of about 136,000, attained by May 2021 (Figure 4 (b), blue curve). Mild social-distancing decreases the peak daily cases and cumulative mortality by 24% and 7%, respectively (Figures 4 (c) and (d), red curve), in relation to the baseline worst-case scenario. Furthermore, moderate social-distancing reduces the peak daily cases and cumulative mortality by 69% and 33% (Figures 4 (c) and (d)), respectively, in comparison to the worst-case scenario. Here, too, the pandemic will fail to take off if strict social-distancing was implemented (and maintained) during the early stages of the pandemic.

For Kano State, the peak daily cases for the worst-case scenario (of about 8,800) is attained around June 1, 2020 (Figure 4 (e)). This corresponds to a peak cumulative mortality of about 276,500, attained around July 2021 (Figure 4 (f)). While mild social-distancing reduces the peak and cumulative mortality by 14% and 3%, in comparison to the worst-case scenario, the implementation of moderate social-distancing in the state reduces these numbers by 47% and 12%. The implementation of strict social-distancing in the state reduces these numbers by 84% and 42%, respectively. Our simulations show that strict social-distancing may not be enough to prevent the pandemic from taking off in Kano State.

Finally, for the Federal Capital Territory, simulations depicted in Figures 4 (g) and (f) show that the peak daily cases for the worst-case scenario (of about 3,000) is achieved around August 2020 (Figure 4 (g), blue curve). This corresponds to a cumulative mortality of 174,100, attained around October 2021 (Figure 4 (h), blue curve). When mild social-distancing measures are implemented, the peak daily cases shifts to October 2020, and 23% and 8% reductions in the peak daily cases and cumulative mortality are, respectively, achieved (Figures 4 (g) and (h), red curve). Although moderate social-distancing may not be adequate to prevent the pandemic from taking off in the FCT, the implementation of strict social-distancing from the onset of the COVID-19 pandemic would have effectively suppressed the pandemic from taking off in the FCT.

### 4.2 Effect of Wearing Face Masks in Public

The community-wide effect of using face masks in public is monitored by simulating the model (2.1), using the baseline parameters tabulated in Tables 3 and 4, using various levels of face mask compliance and efficacy. For these simulations, social-distancing is implemented at the baseline level (given by the baseline values of the community contact rate parameters, *β*_*s*_ and *β*_*a*_, given in Table 3). Specifically, contour plots of the control reproduction number (ℛ_*c*_), as a function of face mask efficacy (*ε*_*M*_) and compliance (*c*_*M*_), are generated for the four communities (Figure 5). These contours show a decrease in the control reproduction number with increasing face mask efficacy and compliance, as expected. In particular, Figure 5 (a) shows that although the use of face masks, with the assumed baseline face mask efficacy of 50%, in Nigeria, Lagos and Kano States can reduce the COVID-19 burden (by reducing ℛ_*c*_), it fails to reduce ℛ_*c*_ to a value less than unity (which is needed for the elimination of the pandemic, in line with Theorem 3.1) even if everyone wears mask in public. However, COVID-19 can be eliminated in the FCT, if at least 96% of the residents of the community wear face masks in public. Thus, this study shows that COVID-19 can be effectively controlled in Nigeria using face masks of moderate efficacy (e.g., face masks with 50% efficacy) if the compliance in their usage is high enough (in the extremely high range of 80% to 95%). These face mask compliance figures (of 80% to 95%) are certainly not realistically-attainable in Nigeria, or anywhere else in the world. Hence, the control strategy based on the use of face masks (with moderate efficacy) in the public should be complemented with other non-pharmaceutical interventions, such as social-distancing and community lockdown, to achieve realistic effective control of the pandemic in Nigeria.

**Figure 5:**
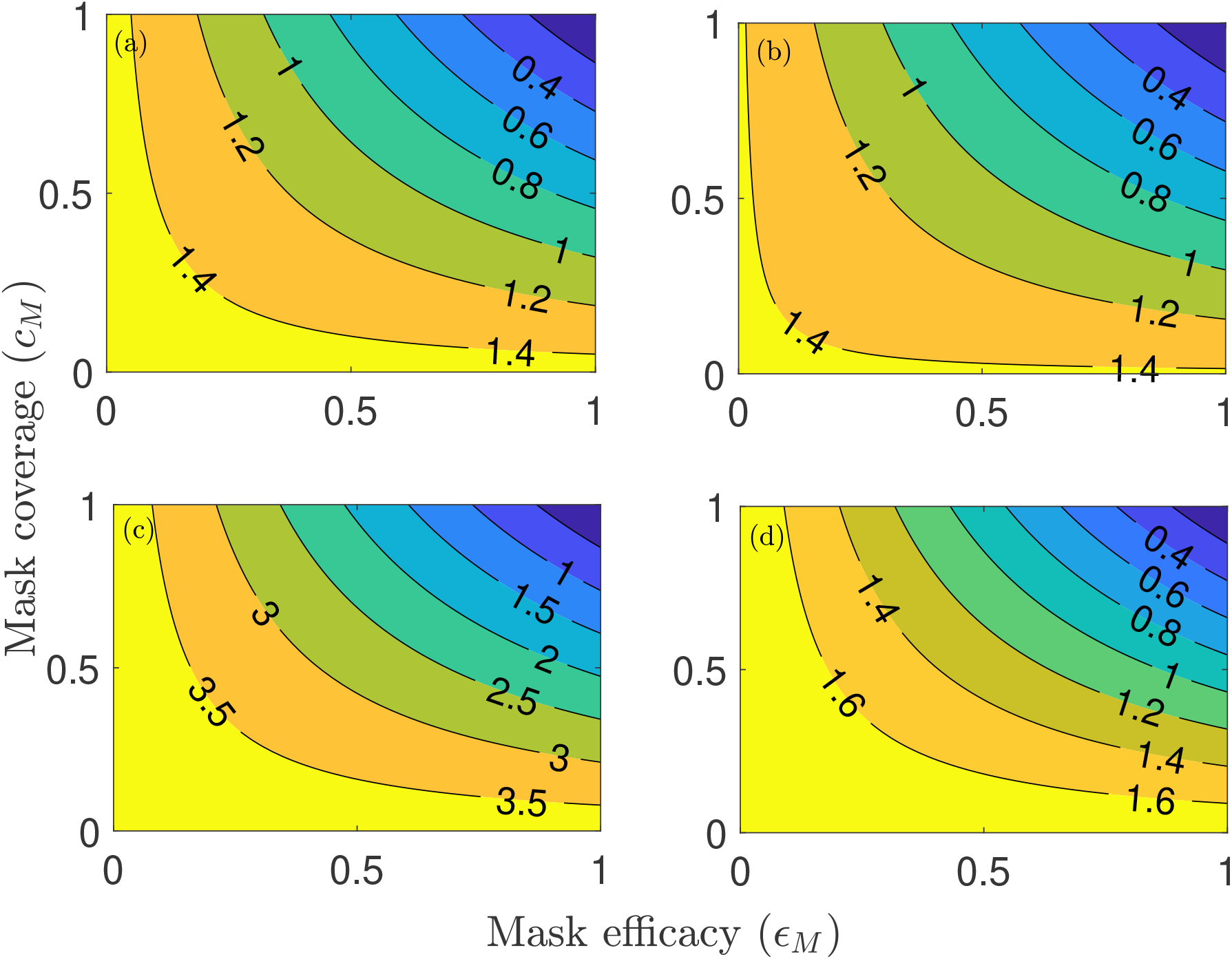
Contour plots of the control reproduction number (ℛ_*c*_) of the model (2.1), as a function of mask coverage (*c*_*M*_) and mask efficacy (*ε*_*M*_), for the special case of the model with no social-distancing. (a) the entire Nigerian nation. (b) Lagos State. (c) Kano State. (d) FCT Abuja. Parameter values used are as given in Tables 3 and 4.

Figure 6 depicts the cumulative mortality recorded in the four communities with various efficacy and coverage levels of face masks. For low quality face masks (such as home-made cloth masks, with estimated efficacy of about 25%), our simulations show that, for the worst-case scenario where masks are not used in public, up to 225,000 deaths can be recorded in Nigeria by February 2021 (Figure 6 (a)). This number decreases, of course, when face masks are used in public, and with increasing compliance. For instance, the cumulative mortality associated with the use of face masks of the low efficacy of 25% reduces to 210,000 (representing 7% reduction, in comparison to the worst-case scenario with no mask usage) if 50% of the Nigerian populace wear face masks in public. The reduction increases to 11% if 75% of Nigerians wear face masks in public. Similar reductions are observed in the States of Lagos (Figure 6 (b)), Kano (Figure 6 (c)) and the FCT (Figure 6 (d)). Thus, as noted by Eikenberry *et al*. [4] and Ngonghala *et al*. [5], the use of face masks in public, even those with low efficacy (such as the home-made masks with efficacy of about 25%), is useful. However, their impact in reducing the cumulative COVID-19 mortality is somewhat marginal (since it only resulted in 8% to 14% reduction in the cumulative mortality, in relation to the worst-case scenario).

**Figure 6:**
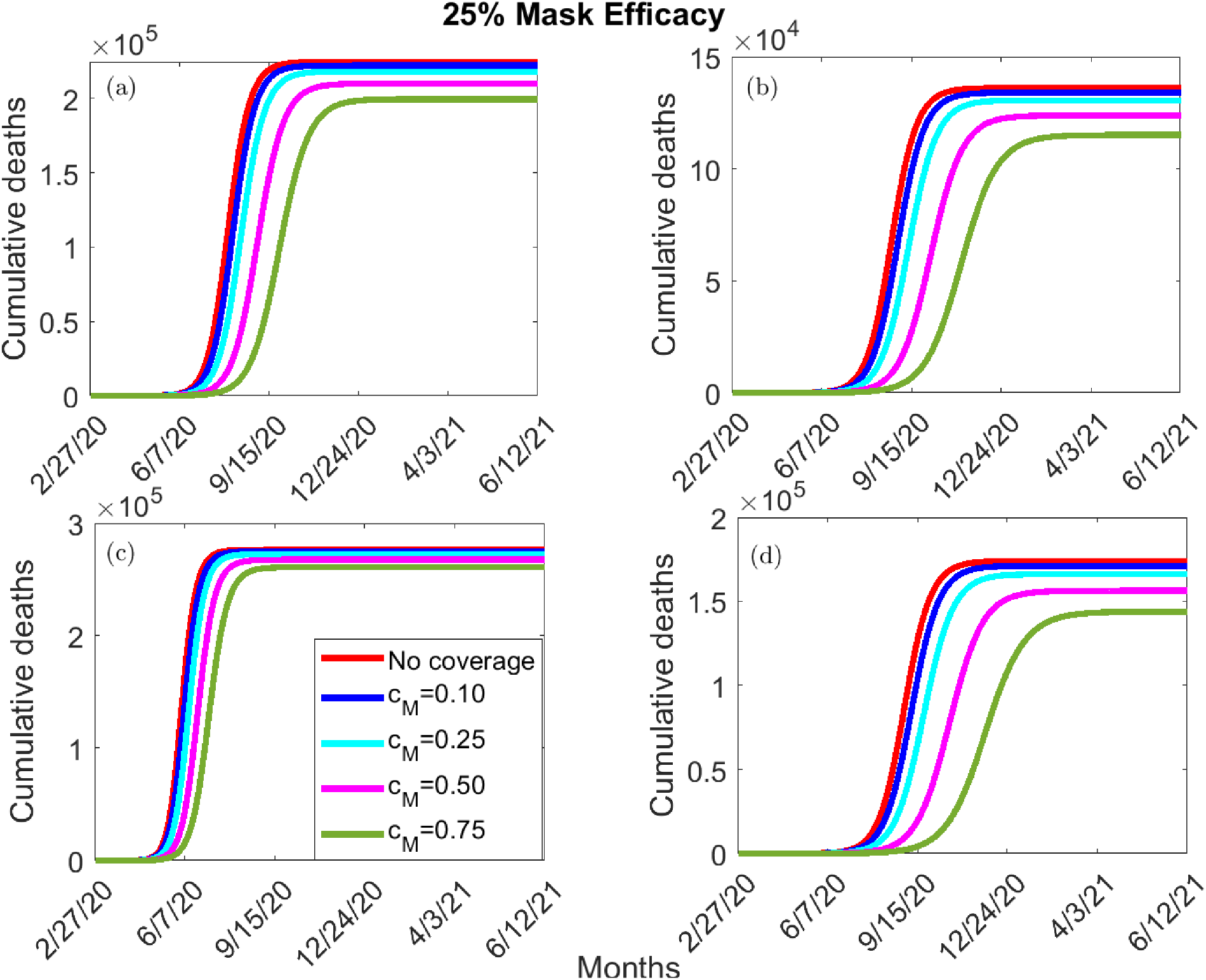
Simulations of the model (2.1), showing cumulative deaths, as a function of time, for 25% face-mask efficacy (*ϵ*_*M*_) and coverage (*c*_*M*_). (a) the entire Nigerian nation. (b) Lagos State. (c) Kano State. (d) FCT Abuja. Parameter values used are as given in Tables 3 and 4.

If masks of moderate efficacy are used (such as masks of efficacy of 50%), our simulations show a more dramatic reduction in the cumulative mortality in the four communities with increasing masks compliance (Figure 7). In particular, Figure 7 (a) shows that, while up to 225,000 deaths could be recorded in Nigeria by February 2021 under the worst-case scenario (Figure 7 (a), red curve), the cumulative deaths can decrease to as low as 141,000 by the same time if 75% of Nigerians wear the moderately-effective face masks in public (Figure 7 (a), green curve). This represents a whopping 37% reduction (in cumulative mortality) from the worst-case scenario baseline. For Lagos State, our simulations show that the use of the moderately-effective (i.e., 50% efficacious) face mask can lead to a reduction of up to 24% if 50% of Lagosians wear face masks in public (Figure 7 (b), purple curve). Further, there is a reduction of up to 81% if 75% of Lagosians wear the moderately-effective face mask in public (Figure 7 (b), green curve). Similar results are obtained for the COVID-19 dynamics in Kano State (Figure 7 (c)) and the FCT (Figure 7 (d)). Even more dramatic reductions in COVID-19 burden in the four communities are recorded if face masks of very high efficacy (e.g., masks of 75% efficacy, such as the N95 respirators) are used (Figure 8). Thus, in summary, these simulations show that although the use of face masks in public can significantly decrease COVID-19 burden in Nigeria (if the face mask efficacy and compliance are high enough), it may fail to eliminate the disease nationwide. However, such elimination is feasible in Kano and Lagos States, as well as in the FCT, if residents wear face masks in public even if the efficacy of the face masks, and compliance in their usage, are moderate.

**Figure 7:**
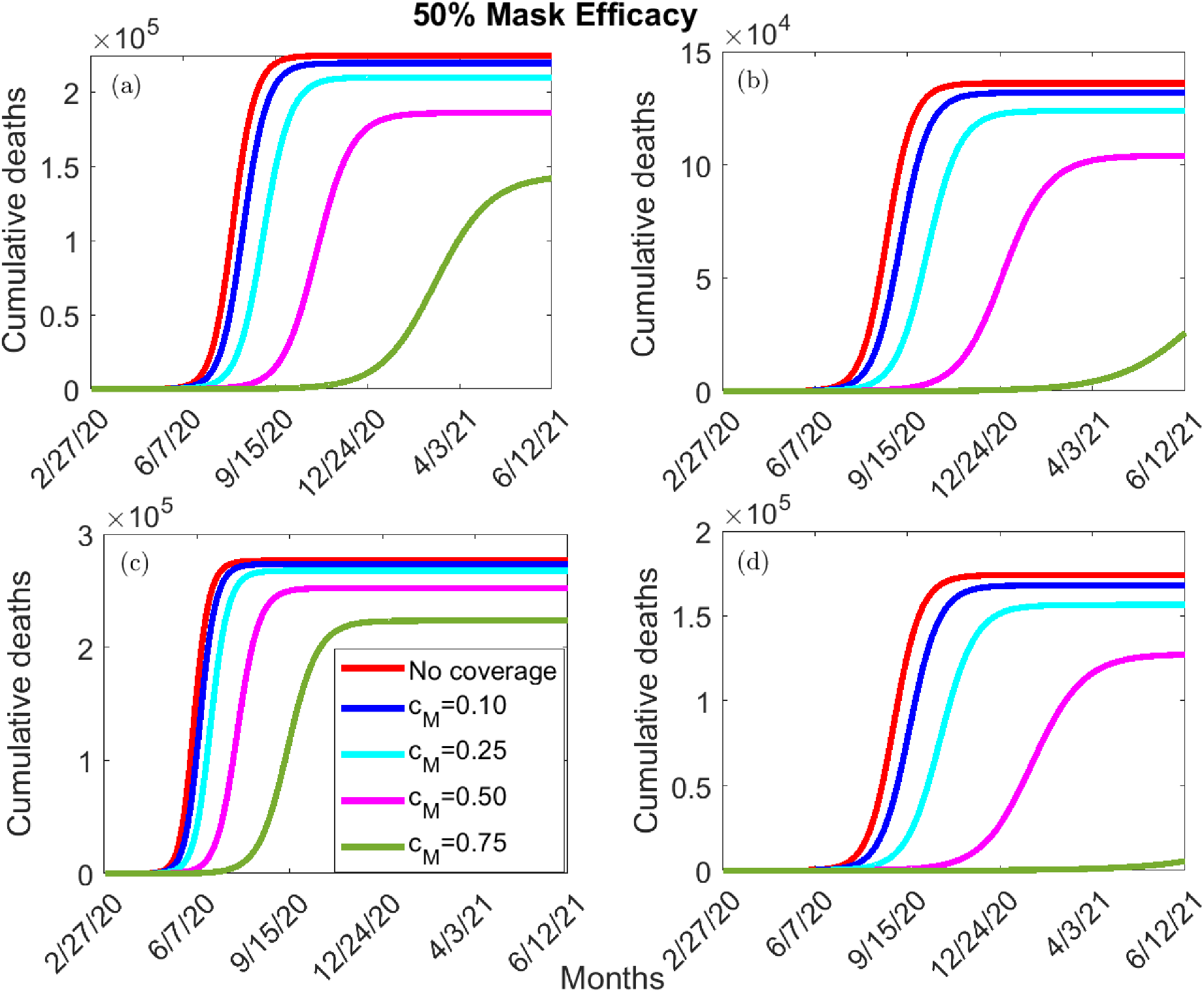
Simulations of the model (2.1), showing cumulative deaths, as a function of time, for 50% face-mask efficacy (ϵ_*M*_) and coverage (*c*_*M*_). (a) the entire Nigeria. (b) Lagos State. (c) Kano State. (d) FCT Abuja. Parameter values used are as given in Tables 3 and 4.

**Figure 8:**
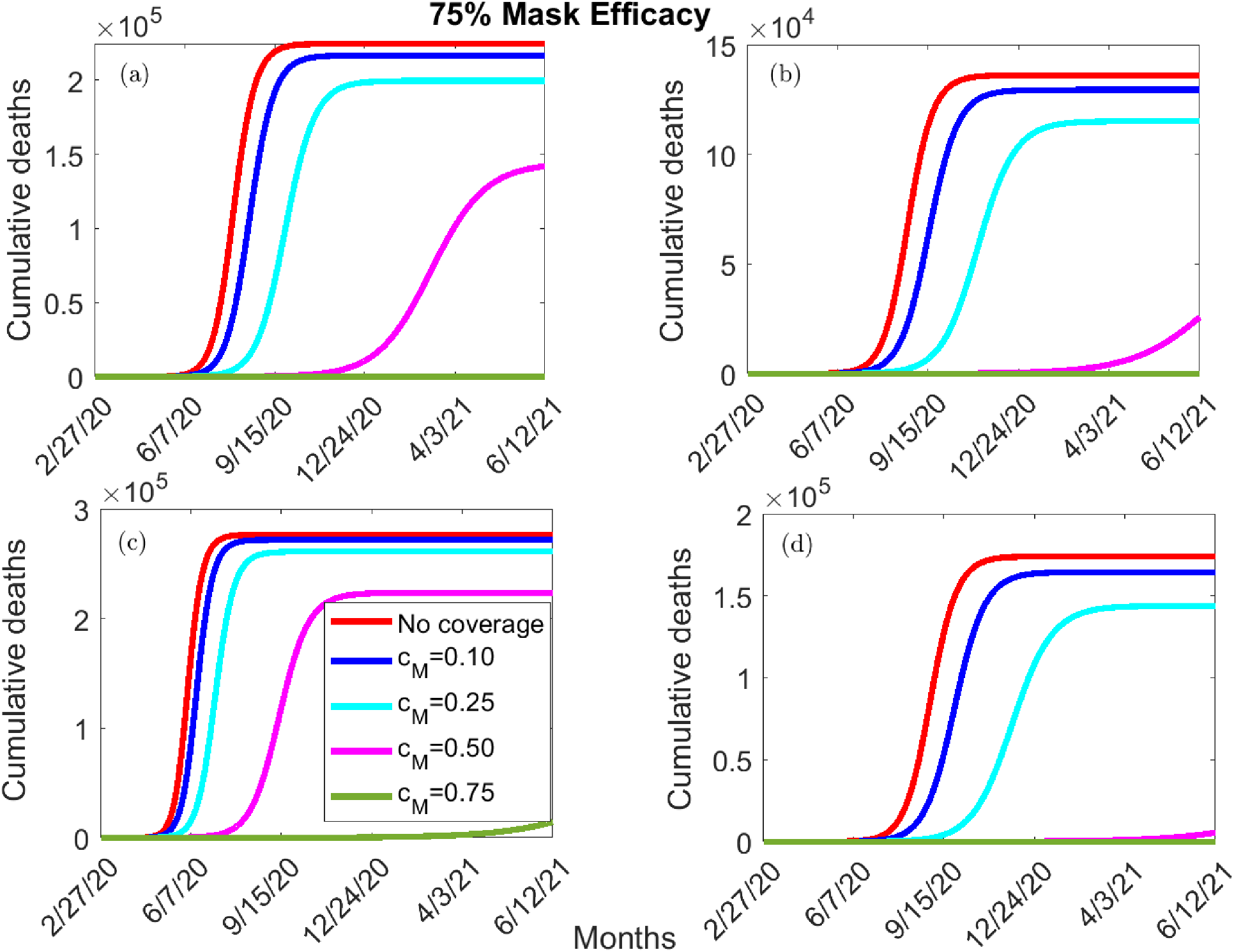
Simulations of the model (2.1), showing cumulative deaths, as a function of time, for 75% face-mask efficacy (ϵ_*M*_) and coverage (*c*_*M*_). (a) the entire Nigeria. (b) Lagos State. (c) Kano State. (d) FCT Abuja. Parameter values used are as given in Tables 3 and 4.

### 4.3 Effect of Early Relaxation/Lifting of Community Lockdown Measures

The community-wide impact of the length (or duration) of the community lockdown measures implemented in Nigeria is monitored by simulating the model (2.1), using the baseline parameter values in Tables 3 and 4 in the absence of face mask usage in public, for various time duration of the community lockdown. It should be recalled that the index case was reported in Nigeria on February 27, 2020, and lockdown measures were initially announced in Nigeria on March 30, 2020. The purpose of these simulations is to determine when it would be safe to consider easing or lifting the lockdown measures in Nigeria. In other words, when can we consider any kind of lifting or easing of the strict lockdown currently in place without triggering the possibility of a second wave of the pandemic that may be as (or even more) devastating as the first wave. For these simulations, we consider the following three levels of lifting of lockdown measures:

i. **Mild lifting (easing) of lockdown measures**: this entails increasing the community contact rates (*β*_*s*_ and *β*_*a*_) by 10% from their baseline values in Table 3.
ii. **Moderate lifting (easing) of lockdown measures**: this entails increasing the community contact rates (*β*_*s*_ and *β*_*a*_) by 30% from their baseline values in Table 3.
iii. **Full lifting (easing) of lockdown measures**: this entails increasing the community contact rates (*β*_*s*_ and *β*_*a*_) by 70% from their baseline values in Table 3.

We first simulated the case when the lockdown measures are lifted/eased after a month of their implementation. That is, we simulated the case when lockdown measures are eased/lifted on April 30, 2020. The results obtained, depicted in Figure 9, showed that the cumulative mortality increases with increasing level of the lifting/easing allowed. In particular, in the case when no social-distancing and lockdown measures are implemented in the country (i.e., when we simulate the worst-case scenario of the model with the baseline values of the community contact rates in Table 3), Nigeria will attain a peak cumulative mortality of about 225,000 by February 2021. Figure 9 (a) shows that when the lifting is mild, up to 232,400 cumulative deaths will be recorded by December 2020 (Figure 9 (a), blue curve). This exceeds the aforementioned cumulative mortality for the worst-case scenario (of 225,000 deaths). If the lifting is moderate or full, the projected cumulative deaths increased (from the worst-case scenario baseline) by 8% and 11%, respectively. This, this study shows that lifting/easing the lockdown in Nigeria after a month of its implementation will trigger a second wave that will be more devastating (8% to 11%) than the first.

**Figure 9:**
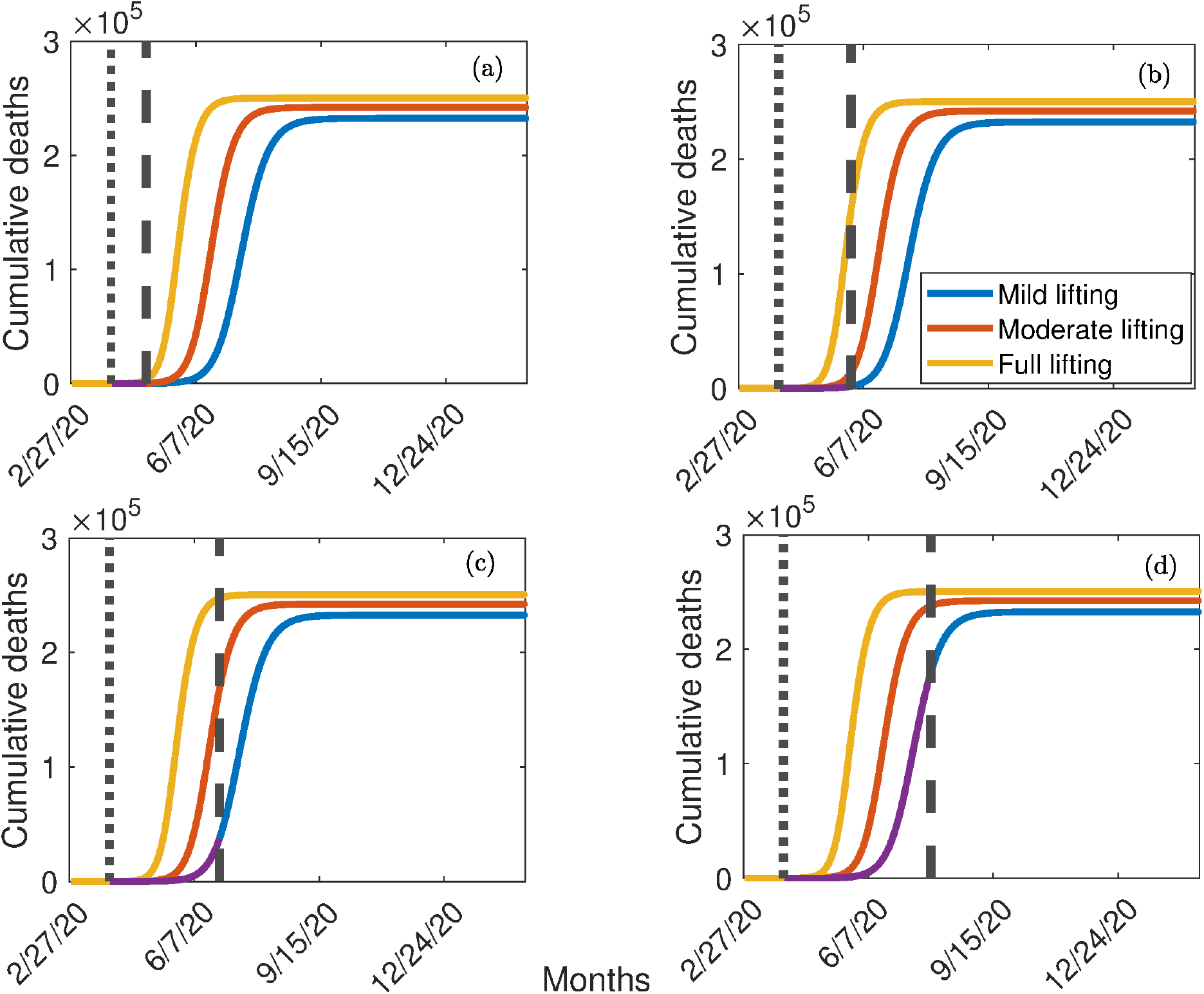
Simulations of the model (2.1), showing cumulative deaths, as a function of time in the absence of mask usage, for the entire Nigerian nation. (a) one month after implementing the lockdown on March 30, 2020 (b) two months after implementing the lockdown on March 30, 2020 (c) three months after implementing the lockdown on March 30, 2020 (d) four months after implementing the lockdown on March 30, 2020. The solid dotted vertical lines indicate the start of the social-distancing lockdown regimen (March 30, 2020), dashed vertical lines indicate when the current strict social-distancing is terminated, and the purple horizontal line segments indicate the duration of the strict social-distancing regimen. Parameter values used are as given in Tables 3 and 4.

When the lockdown measures are lifted after two months of their implementation (on May 30, 2020), our simulations show up to 231,200 cumulative deaths will be recorded by December 2020 if the lifting is mild (Figure 9 (b)). Again, this exceeds the cumulative mortality recorded under the worst-case scenario. Furthermore, if the lifting is moderate or full, the projected cumulative deaths increased by 7% and 11%, respectively. When the lockdown measures are lifted/eased after three months of implementation (on June 30, 2020), our simulations show that mild lifting will generate up to 188,500 cumulative deaths by December 2020 (Figure 9 (c)). This represent a 16% decrease in the worst-case scenario projection (of 225,000 deaths). Thus, running the lockdown measures for at least three months period will significantly lower the burden of the pandemic in Nigeria. Furthermore, if the lifting is moderate or full, the projected cumulative deaths decreased by 12% and 8%, respectively, from the worst-case scenario baseline. Finally, when the lockdown measures are lifted after four months of their implementation (on July 30, 2020), our simulations show that up to 37,500 cumulative deaths will be recorded by December 2020 if the lifting/easing is mild (Figure 9 (d)). This represents a 83% decrease from the worst-case scenario baseline. If the lifting is moderate or full, the projected cumulative deaths decreased by 79% and 75%, respectively, from the worst-case scenario baseline. Thus, this study shows that extending the lockdown to at least four months will greatly help in curtailing the COVID-19 pandemic in Nigeria. In fact, our study shows that if the lockdown measures are extended until the end of 2020, the projected cumulative deaths decreases by 100%, 96% and 92% if the lifting is mild, moderate and full. In other words, we can actually eliminate the pandemic if the lockdown measures are extended until the end of the year. Similar results are obtained for Lagos State, Kano State and the FCT (not reported here to save space). In summary, this study shows that running the lockdown measures to longer periods (at least four months) will greatly enhance the prospects of effectively curtailing or eliminated the pandemic. However, the early lifting or relation of the social-distancing and lockdown measures, in an effort to re-open the economy and the country, could trigger a second wave of the COVID-19 pandemic that may be more devastating than the first wave.

## Discussion and Conclusions

The world has been facing a devastating novel Coronavirus pandemic (COVID-19), caused by SARS-CoV-2, since its emergence in the Wuhan city of China at the end of 2019. The pandemic, which has rapidly spread to over 210 countries, continues to inflict severe public health and socio-economic burden in many parts of the world, including in Nigeria. It has, as of May 18, 2020, accounted for over 4.7 million confirmed cases and about 315,000 deaths globally. In the absence of a safe and effective vaccine or antiviral for use against the pandemic in humans, control efforts are focused on the use of non-pharmaceutical interventions (NPIs), such as social (physical)-distancing, community lockdown, contact tracing, quarantine of suspected cases, isolation of confirmed cases and the use of face masks in public. Nigeria is one of the countries in Africa that is hardest hit with the burden of the COVID-19 pandemic. We developed a mathematical model for assessing the COVID-19 pandemic in Nigeria, and use the model to assess the community-wide impact of the various control and mitigation strategies implemented nation-wide, as well in two states of the Nigerian federation (Kano and Lagos) and the Federal Capital Territory (FCT Abuja), to effectively curtail the pandemic. The model was parametrized using COVID-19 data published by the Nigeria Centre for Disease Control (NCDC).

The Kermack-McKendrick compartmental epidemic model we developed in this study, which takes the form of a deterministic system of nonlinear differential equations, has a continuum of disease-free equilibria, which is rigorously shown to be locally-asymptotically stable whenever a certain epidemiological threshold, known as the control reproduction number (denoted by ℛ_*c*_), is less than unity. The epidemiological implication of this result is that the COVID-19 pandemic can be effectively controlled in Nigeria, whenever ℛ_*c*_ *<* 1, if the initial sub-populations of the infected compartments of the model are small enough. In other words, COVID-19 can be effectively controlled in Nigeria if the control and mitigation strategies implemented are able to bring (and maintain) the threshold quantity (ℛ_*c*_) to a value less than unity. An expression for the final size of the pandemic was also theoretically derived (which is expressed in terms of the basic reproduction number (ℛ_0_) of the model and the number of individuals in the community that remain in each of the disease-free compartments of the model (namely, those in the susceptible and recovered compartments) during the course of the COVID-19 pandemic in Nigeria).

After parameterizing the model, to realistically estimate some of the pertinent missing parameters (such as the community contact rate parameters), we carried out numerous numerical simulations of the model to assess the effectiveness of the various control strategies implemented in Nigeria in general, and in the three other communities (Kano and Lagos States, and the FCT Abuja) in particular. We simulated the impact of social-distancing in curtailing the impact of COVID-19 in Nigeria (as a whole) first of all. We considered three effectiveness levels of social-distancing, namely mild, moderate and strict (depending on the percent reduction in the associated community contact rate parameters, *β*_*s*_ and *β*_*a*_). Our simulations showed that the implementation of mild social-distancing measures can significantly decrease COVID-19 burden in Nigeria (as measured in terms of shifting and lowering the peak daily cases and cumulative mortality nationwide). Although a further (more dramatic) reduction in the pandemic burden is achieved nationwide if the social-distancing measures are implemented at moderate level of effectiveness, outbreaks of the COVID-19 pandemic will continue to be occurring (i.e., the pandemic will continue to be spreading, *albeit* at a much slower rate, in comparison to when low level or no social-distancing measures are implemented). On the other hand, if strict social-distancing measures are implemented, the pandemic will actually fail to effectively take off at all in the country. In other words, our study shows that if strict social-distancing measures are implemented (and maintained for an extended period of time), COVID-19 will be unable to cause significant outbreaks in Nigeria. Similar results were obtained when such effectiveness levels of social-distancing measures are implemented in Kano and Lagos States. Our study further showed that, unlike in the entire Nigerian nation and the states of Lagos and Kano (where the COVID-19 pandemic causes outbreaks even if social-distancing measures with moderate effectiveness level are implemented), the COVID-19 will not be able to cause significant outbreaks in the Federal Capital Territory if moderate social-distancing measures are implemented. This result is intuitive in the sense that the Federal Capital Territory is not as highly-dense as the rest of the country in general, and certainly not as dense as the highly-populated Kano and Lagos States.

The use of face masks (including home-made cloths masks and medical masks) has been strongly advocated by public health officials around the world, as a vital tool to help minimize community transmission of COVID-These masks have inward and outward efficacies to limit both the acquisition of, as well as the transmission of, COVID-19 infection. We simulated our model to monitor the community-wide impact of using face masks in public in Nigeria. Our study showed a decrease in COVID-19 burden, as measured in terms of the decrease in the control reproduction number of the model (ℛ_*c*_), with increasing face mask efficacy and compliance, as expected. In particular, we showed that COVID-19 can be eliminated in Kano and Lagos States, as well as in the FCT, if the overwhelming majority of the population (between 80% to 95%) consistently wear a moderately-efficacious (of efficacy 50%) in public. This mask compliance proportions are certainly not realistically feasible in Nigeria, or in any other part of the world. Hence, while the use of face masks of moderate efficacy can significantly reduce the burden of the pandemic, their ability to lead to the elimination of the pandemic requires unrealistic compliance level. Hence, although the use of moderately-efficacious face masks in public is always useful, masks alone cannot realistically lead to the elimination of the pandemic. To achieve such elimination using moderately-efficacious face masks, the face masks-based strategy needs to be complemented with other non-pharmaceutical strategies that aim to reduce community transmission (such as social-distancing and community lockdown, contact tracing etc.). When highly efficacious masks (such as the N95 respirators) in public, our study showed that COVID-19 can, indeed, be eliminated in Nigeria. These highly-efficacious face masks are in short supply, and are (rightly) reserved for the frontline health care workers. Hence, the use of these highly-efficacious masks is not a realistic option. In summary, our study showed that, although the use of moderately-efficacious face masks in public can significantly decrease COVID-19 burden in Nigeria, it will be inadequate to eliminate the disease nationwide. However, such elimination is realistically-achievable in Kano and Lagos States, as well as in the FCT, using the moderately-efficatious masks in public if the compliance in their usage is at least moderate.

Nigeria implemented community lockdown measures on March 30, 2020. Although the lockdown measures have proved to be effective in reducing the burden on the pandemic, a large number of the citizenry expressed concern and distress caused by the lockdown (as is generally the case in every community in the rest of the world that implemented such lockdowns). There is now strong clamour to ease or fully lift the lockdown in Nigeria. The main challenge is, of course, to ensure that any easing or lifting of the lockdown measures does not erase the gains made in curtailing the pandemic during the lockdown. In other words, utmost care must be taken before easing/lifting the lockdown to avoid triggering a second wave of the pandemic (that could be as, or more, disastrous as the first). We simulated our model to determine the safe time for such measures to be be eased in Nigeria. Our simulations showed that the lockdown measures should be in place for at least three to four months (from the March 30, 2020 initial implementation date) to ensure that the possibility of a devastating second wave is minimized. We showed an even more promising result (of completely curtailing the pandemic outbreak in Nigeria) if the existing lockdown measures are maintained until December 2020. Relaxing or terminating the lockdown measures too soon, as part of the move to re-open the economy and the country, would undoubtedly trigger a deadly rebound of COVID-19 in Nigeria.

Community lockdown surely induces severe burden and stress on the people. Nonetheless, it is a necessary measure to take to avert catastrophic loss of life. Consequently, governments at all levels (federal, state and local) have a responsibility to do everything they can to alleviate the sufferings of the people during lockdown. In particular, effective measures such as the injection of sizable stimulus package into the economy, the provision of food shelters, curtailing the gouging of prices of commodities and essential goods and services, helping employers to pay their employees (*via* strategic plans, such as Paycheck Protection Program) etc. Citizens support is crucially needed for the lockdown program to be successful. The city of Wuhan was in lockdown for 76 days. The city of New York has been on stay-at-home lockdown since March 22, 2020, and the lockdown is expected to stay in place until at least June, 2020 (it should be mentioned that the State of New York issued an executive order making it mandatory for people in New York city to wear face masks in public [29]).

Our projections for COVID-19 burden in Nigeria seems to be significantly higher than what would have reasonably been expected from the data reported by the Nigeria Centre for Disease Control (NCDC). We believe that the data reported by the NCDC may represent significant under-estimation of the true burden of the pandemic in Nigeria. There are many factors to help substantiate this claim. First, even with the seemingly flawed data reported by the NCDC, our estimate of the basic reproduction number of the model (ℛ_0_) for Nigeria (and any of the three other population centres within Nigeria we considered) is ℛ_0_ *≈* 2. That is, on average, one COVID-infected person in Nigeria transmits to two other Nigerians. Hence, from the reported date the index case was diagnosed (February 27, 2020) until control measures were implemented (March 30, 2020), the pandemic was spreading in Nigeria unabated. and at this ferocious exponential speed! Hence, it stands to reason that a large number of Nigerians have already acquired (and equally a large number have succumbed to) the pandemic before control measures were implemented. Sadly, all these cases (and deaths) that were occurring before the control measures were put in place would obviously not have been recorded in the NCDC databases. Further, the fact that not much COVID-19 diagnostic testing was done in Nigeria (only about 35,345 people were tested, in a country of 200 million people, as of May 18, 2020 [9]). This, together with the fact that autopsies and testing of deceased individuals are not generally (or universally) carried out in the country (in some cases, for traditional and religious reasons), clearly suggests a gross under-reporting of the true burden of the pandemic. It is also intuitive to assume that the pandemic was already spreading in Nigeria way before the index case was reported on February 27, 2020. This is owing to the huge volumes of traffic between Nigeria and China (the first epicenter of COVID-19) and Europe (the second epicenter of COVID-19) during the last few months of 2019 and the early months of 2020.

Taking the aforementioned factors and hypotheses in totality, it seems quite plausible to support the notion that the NCDC data grossly under-estimated the burden of the pandemic in Nigeria, and that the projections of realistically-formulated mathematical models, such as ours, may well provide a more reasonable estimate of the true burden of the COVID-19 pandemic in Nigeria, than the data provided by the NCDC. Nigeria’s large population size (of about 200 million) and volume of air traffic, to and from the rest of the world, coupled with the high population densities in its major cities (such as Kano and Lagos), makes it ripe to suffer devastating COVID-19 burden on the scale similar to what transpired in the US state of New York (with its major population centres having similar population size and density, and also being major hubs for global travel, particularly, to and from, Europe). Unless effective public health interventions are implemented, and sustained for a significant period of time (as was done, and is being done, in New York state), Nigeria would undoubtedly continue to record catastrophic burden of the pandemic (even if the ensuing case and mortality numbers are not realistically recorded by the Nigeria’s public health authorities). However, our study emphasize the fact that the COVID-19 pandemic is controllable in Nigeria using basic non-pharmaceutical interventions, such as social (physical)-distancing, community lockdown, the use of face masks in public, the use of personal protection equipment (PPE) by frontline healthcare workers, widespread diagnostic testing and contact tracing, personal hygiene and hand washing, rapid isolation of confirmed cases and the quarantine of people suspected of being exposed to the pandemic (provided these interventions are implemented at moderate levels of effectiveness and coverage).

It is imperative that Nigeria ramps up widespread diagnostic COVID-19 testing and contact tracing to have a realistic measure of the burden of the pandemic. It is also imperative that Nigeria emphasize personal hygiene and hand washing (using soaps and approved hand sanitizers), physical-distancing, wearing face masks in public and make PPEs widely available to frontline healthcare workers. Re-opening and/or relaxation of the current lockdown measures must be done in a very responsible manner, and based solely on what the data (from the robust widespread testing) says. These measures are necessarily needed to effectively control the spread, and mitigate the burden, of the pandemic, before a safe and effective vaccine or antiviral becomes available. Further, adequate measures must be taken to alleviate the sufferings of citizens under COVID-19 lockdowns (particularly in areas of food security and the supply of other basic amenities of life).

## Data Availability

N/A

## Acknowledgments

One of the authors (ABG) acknowledge the support, in part, of the Simons Foundation (Award #585022) and the National Science Foundation (Award #1917512). CNN acknowledges the support of the Simons Foundation (Award #627346).

## Appendix A: Proof of Theorem 3.3

*Proof*. Consider the model (2.1) with *ℛ*_*c*_ *<* 1. Furthermore, consider the following Lyapunov function:

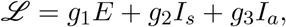

where, *g*_1_ = *γ*_*a*_(*ϕ*_*s*_ + *γ*_*s*_ + *δ*_*s*_), *g*_2_ = *γ*_*a*_*β*_*s*_(1 − *ε*_*M*_ *c*_*M*_), and g_3_ = (*ϕ*_s_ + *γ*_s_ + *δ*_s_)*β*_s_(1 *− ε*_M_c_M_). It follows that the Lyapunov derivative is given by:

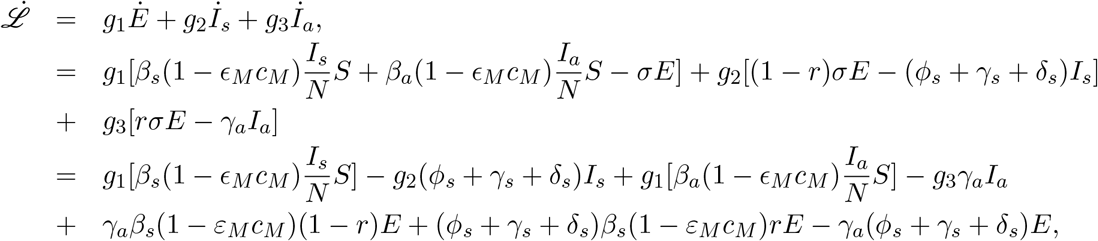

which can be simplified to,

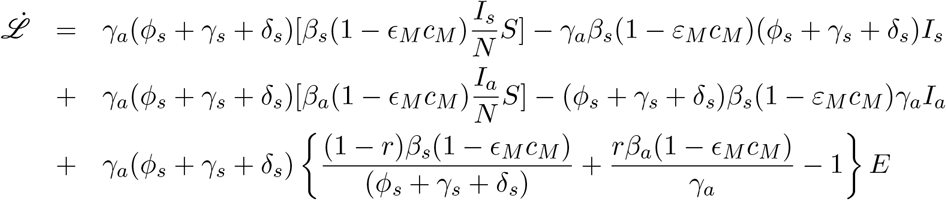

so that (noting that *S*(*t*) *≤ N* (*t*) for all *t* in Ω),

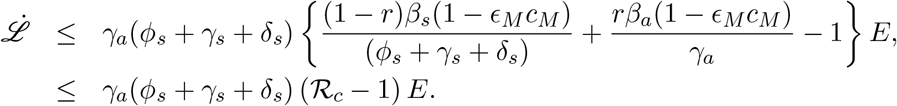

Hence, 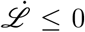 if *ℛ*_*c*_ *≤* 1, and 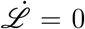 if and only if *E*(*t*) = 0. Substituting *E*(*t*) = 0 in the model (2.1) shows that (*S*(*t*), *E*(*t*), *I*_*s*_(*t*), *I*_*a*_(*t*), *I*_*h*_(*t*), *R*(*t*)) *→* (*N* (0) *− R*^*∗*^, 0, 0, 0, 0, *R*^*∗*^), as *t → ∞*. Further, it can be shown that the largest compact invariant set in {(*S*(*t*), *E*(*t*), *I*_*s*_(*t*), *I*_*a*_(*t*), *I*_*h*_(*t*), *R*(*t*)) *∈* Ω: 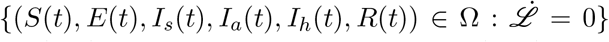 is the line of disease-free equilibria (*ε*_0_). Hence, it follows, by the LaSalle’s Invariance Principle, that the continuum of disease-free equilibria of the model (2.1) is globally-asymptotically stable in Ω whenever *ℛ*_*c*_ *≤* 1.

## Notes

### Competing Interest Statement

The authors have declared no competing interest.

